# Stigma and unsupportive disclosure reactions are associated with elevated depressive symptom burden among adolescent mothers in Juja Sub-County, Kenya: a community-based cross-sectional study

**DOI:** 10.64898/2026.11.17.26368357

**Authors:** Teresia Wanjiru Kariuki, John Kariuki, Dominic Mogere, Jared Onyancha, Gervason Moriasi

## Abstract

Girls who become pregnant during adolescence in sub-Saharan Africa carry a high burden of depressive symptoms. Kenyan studies have relied on antenatal clinic samples and have not looked at stigma, the response to pregnancy disclosure and economic circumstances in the same analysis. We assessed how these factors relate to depressive symptom burden among adolescent mothers in Juja Sub-County, Kiambu County. We surveyed 227 girls who had been pregnant or given birth at 10–19 years, drawn by stratified random sampling from a line list of 466 mapped across five wards. Trained assistants administered a structured questionnaire covering background characteristics, pregnancy and disclosure, stigma, and a seven-item depressive symptom checklist. Elevated symptom burden was set in advance at four or more endorsed items. Two logistic regression models examined factors linked to stigma (n = 199) and to elevated symptom burden (n = 184). Of those with parity data, 194 of 197 had already given birth. Most respondents were 17–19 years old (86.8%) and single (69.2%), 92.5% had no income of their own, and 47.1% had left school early. Nine in ten pregnancies (91.2%) were unplanned, and 41.4% of girls met anger, rejection or indifference when they first disclosed. Stigma was reported by 43.2%, elevated symptom burden by 27.1% and thoughts of self-harm by 16.4%. Only 27.3% had attended any support programme. Being single was the one factor associated with stigma (adjusted odds ratio [AOR] 3.50; 95% confidence interval [CI] 1.48–8.31). Elevated symptom burden was associated with stigma (AOR 3.21; 1.54–6.69) and with an unsupportive disclosure reaction (AOR 2.54; 1.19–5.41), and was less likely with each additional year of age (AOR 0.69; 0.49–0.96). Programme attendance showed no association (AOR 0.89; 0.41–1.94). In this peri-urban community, depressive symptoms clustered among adolescent mothers who had been stigmatised or poorly received when they disclosed their pregnancy, and stigma tracked marital status more closely than poverty. Screening with validated tools at postnatal, child-health and antenatal visits, guidance for families and partners on responding to disclosure, and community work on attitudes to unmarried motherhood deserve testing in this setting.

## Introduction

Each year, girls aged 15–19 years in low- and middle-income countries have roughly 21 million pregnancies, and about half of these are unintended [1]. Complications of pregnancy and childbirth rank among the main causes of death in this age group [1]. The psychological toll is also considerable. Pregnant and parenting adolescents commonly report depression, anxiety, isolation and stigma [2, 3], and in sub-Saharan Africa depression is the mental health problem most often documented in this group [3]. Adolescent mental health services in the region have long been under-resourced [2].

Sub-Saharan Africa has a larger share of the world’s adolescent births than any other region [4]. In Kenya, 15% of women aged 15–19 years have been pregnant at least once [5]. Poverty, little schooling, early marriage and low contraceptive use recur as drivers of adolescent pregnancy across comparable settings [6]. Kiambu County sits next to Nairobi and fares relatively well on most socio-economic measures, yet county records still show several thousand adolescent pregnancies a year [7]. Juja Sub-County, which has grown quickly along the Thika Superhighway, carries a heavy share of these, and local reports link them to poverty, peer influence, weak parental guidance and a shortage of youth-friendly services [7].

Kenyan data on depression in this group come almost entirely from antenatal clinics. In two Nairobi primary care facilities, 43.1% of 153 pregnant adolescents scored 10 or more on the Patient Health Questionnaire-9 (PHQ-9) [8]. At an antenatal clinic in Kangemi, also in Nairobi, 32.9% of 176 pregnant adolescents scored 15 or more; recent stressful events, poor social support, HIV infection and younger age were independent predictors, although stigma was not measured [9]. Among 529 pregnant adolescents in Mwanza, Tanzania, 21.0% had depressive symptoms on the Edinburgh Postnatal Depression Scale, and pregnancy-related stigma, partner violence and marital status each showed independent associations [10]. Two gaps stand out. Clinic samples leave out girls who do not attend services, and they tell us little about the months after birth, when many girls are out of school and dependent on family. We also found no Kenyan study that has modelled stigma and the reaction to disclosure together while accounting for economic disadvantage.

We drew on three bodies of theory (Fig. 1). Bronfenbrenner’s ecological model treats development as the product of nested settings [11]. Following it, we grouped our variables into individual circumstances (age, schooling, income), the household (living arrangement, how the first confidant reacted, contact with support programmes) and the wider normative setting, which we indexed by marital status because marriage still governs whether childbearing is seen as legitimate in many Kenyan communities. Goffman described stigma as an attribute that spoils a person’s standing against social expectations [12]; Link and Phelan later framed it as a process of labelling and stereotyping that depends on social power [13]. Cohen and Wills separated two ways in which social support may protect mental health: a direct, main effect, and a buffering effect that operates only under stress [14]. The reaction a girl receives when she first tells someone about her pregnancy is a concrete measure of the support available to her at that moment. We analysed it as a main effect.

**Fig. 1.**
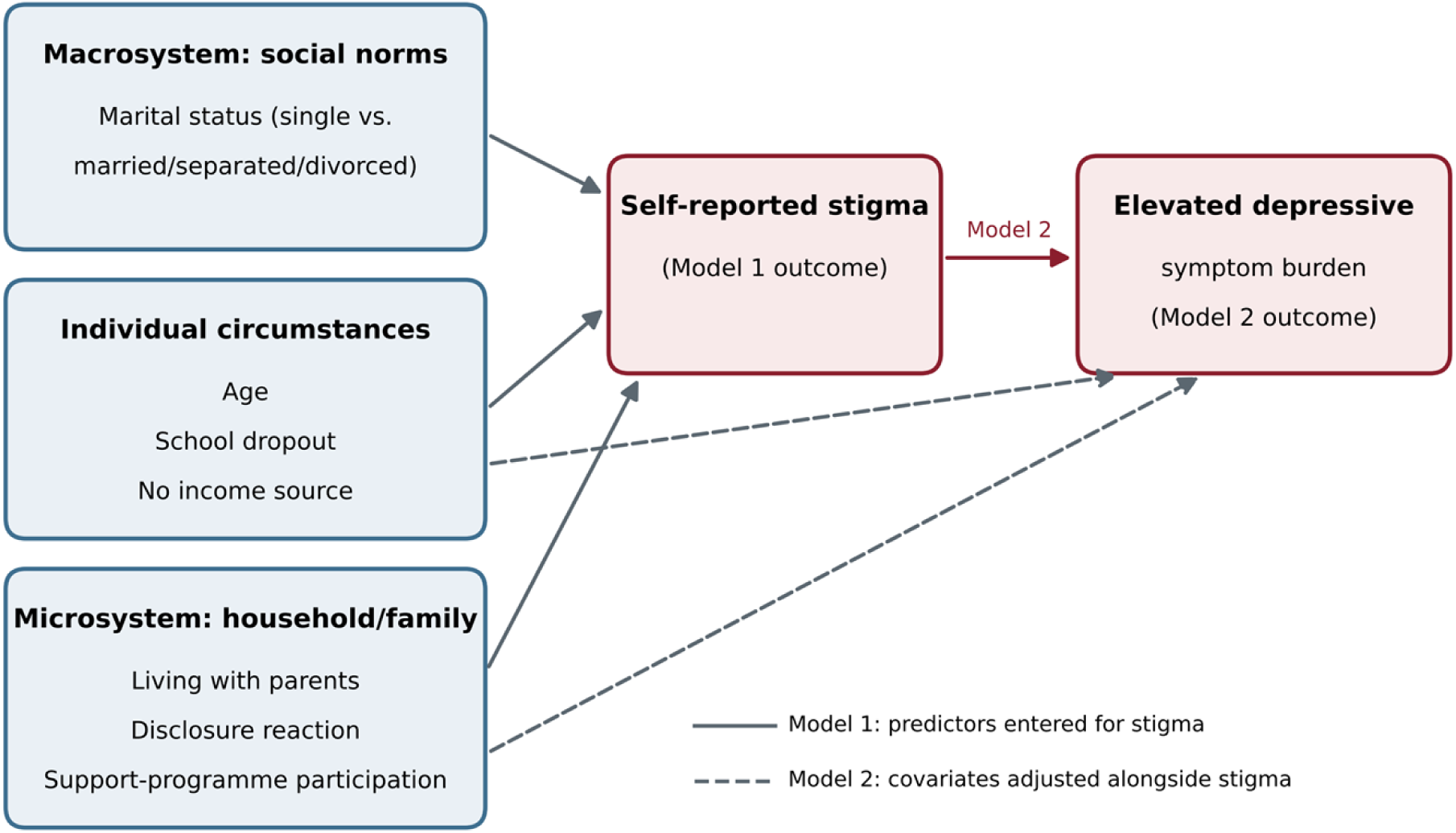
Conceptual framework for the analysis. Variables are grouped by ecological level. Solid grey arrows show the factors entered in the stigma model (Model 1). The red arrow and the dashed arrows show the depressive symptom model (Model 2), in which stigma is entered together with the other covariates. The diagram sets out the analytic structure only; it does not represent tested causal paths.

Work from other African settings points to marital status as a focus of stigma. In a qualitative study across three Nigerian states, adolescent mothers who reported stigma were mostly unmarried and not living with their partners, and a recognised union seemed to protect against it [15]. In Cape Town, norms holding that pregnancy belongs after marriage led adolescents to hide their pregnancies and to expect scolding at clinics [16]. Kenyan nurses have explained poor treatment of pregnant adolescents partly by disapproval of premarital sex [17]. It is not known whether stigma tied to marital status operates in Kiambu, or whether stigma relates to depressive symptoms once disclosure experience and economic hardship are taken into account.

Among pregnant and parenting adolescents in Juja Sub-County, we therefore set out to (i) describe their background, pregnancy and disclosure experiences, stigma and depressive symptoms; (ii) identify factors associated with stigma; and (iii) test whether stigma and the reaction to disclosure are associated with elevated depressive symptom burden after adjustment for economic and educational circumstances. Findings on advocacy programmes, gaps in provision and stakeholder views from the same parent study are reported separately [18].

## Methods

### Study design and setting

This is the quantitative arm of a cross-sectional, community-based study that used a concurrent mixed-methods design. Reporting follows the STROBE statement for cross-sectional studies [19] (Additional file 1). Juja Sub-County has five wards (Witeithie, Kalimoni, Juja, Murera and Theta) and counted 300,948 residents at the 2019 census [20]. The sub-county mixes urban and rural areas, hosts several universities and colleges, and has few mental health services designed for young people [7]. Data were collected from January to July 2026.

### Participants and eligibility

We included girls living in Juja Sub-County who were pregnant, or had given birth, at age 10–19 years. Because eligibility depended on age at the index pregnancy or birth, three respondents were 20 years old at interview; the youngest was 14 (Table 1). Girls who appeared too distressed to take part safely were not interviewed and were referred directly for care, as described under Ethical considerations.

**Table 1.** Characteristics of respondents (N = 227)

| **Characteristic** | **n** | **%** | **95% CI** |
|---|---|---|---|
| **Age (years)** |  |  |  |
| 14–15 | 2 | 0.9 | 0.2–3.2 |
| 16 | 20 | 8.8 | 5.8–13.2 |
| 17 | 52 | 22.9 | 17.9–28.8 |
| 18 | 58 | 25.6 | 20.3–31.6 |
| 19 | 87 | 38.3 | 32.2–44.8 |
| 20<sup>a</sup> | 3 | 1.3 | 0.5–3.8 |
| Missing/invalid<sup>b</sup> | 5 | 2.2 | 0.9–5.1 |
| **Schooling status** |  |  |  |
| Dropped out | 107 | 47.1 | 40.7–53.6 |
| Completed secondary | 56 | 24.7 | 19.5–30.7 |
| Completed primary | 31 | 13.7 | 9.8–18.7 |
| In school | 23 | 10.1 | 6.8–14.7 |
| Never attended | 1 | 0.4 | 0.1–2.5 |
| Missing/unclear | 9 | 4.0 | 2.1–7.4 |
| **Marital status** |  |  |  |
| Single | 157 | 69.2 | 62.9–74.8 |
| Married | 46 | 20.3 | 15.6–26.0 |
| Separated | 15 | 6.6 | 4.0–10.6 |
| Divorced | 1 | 0.4 | 0.1–2.5 |
| Missing/unclear | 8 | 3.5 | 1.8–6.8 |
| **Number of children** |  |  |  |
| None (currently pregnant) | 3 | 1.3 | 0.5–3.8 |
| One | 180 | 79.3 | 73.6–84.1 |
| Two | 14 | 6.2 | 3.7–10.1 |
| Missing/unclear | 30 | 13.2 | 9.4–18.2 |
| **Living arrangement** |  |  |  |
| With parents/guardian | 147 | 64.8 | 58.3–70.7 |
| With partner | 55 | 24.2 | 19.1–30.2 |
| Alone | 13 | 5.7 | 3.4–9.6 |
| In a children’s home | 4 | 1.8 | 0.7–4.4 |
| With friends | 1 | 0.4 | 0.1–2.5 |
| Missing/unclear | 7 | 3.1 | 1.5–6.2 |
| **Main source of income** |  |  |  |
| No income | 88 | 38.8 | 32.7–45.2 |
| Parents | 83 | 36.6 | 30.6–43.0 |
| Partner | 39 | 17.2 | 12.8–22.6 |
| Own job | 8 | 3.5 | 1.8–6.8 |
| Other/mixed | 5 | 2.2 | 0.9–5.1 |
| Missing/unclear | 4 | 1.8 | 0.7–4.4 |
| **Disability or chronic health condition** |  |  |  |
| None reported | 224 | 98.7 | 96.2–99.5 |
| Missing/unclear | 3 | 1.3 | 0.5–3.8 |

CI, confidence interval (Wilson score method). <sup>a</sup>Eligibility was based on age at the index pregnancy or birth. <sup>b</sup>Includes one recorded age of 62 years, treated as an entry error.

### Sample size and sampling

Community health volunteers and health facility records were used to map eligible girls, giving a line list of 466 across the five wards. Cochran’s formula (Z = 1.96, p = 0.5, d = 0.05) gave an initial sample of 384, which the finite population correction for N = 466 reduced to a minimum of 211 [21]. Wards formed the strata. Girls were drawn at random from each ward’s list in proportion to that ward’s share of the mapped total, and recruitment continued beyond the minimum to allow for incomplete questionnaires. The mapped and analysed numbers per ward were: Witeithie 40 and 20; Kalimoni 144 and 70; Juja 115 and 56; Murera 134 and 65; and Theta 33 and 16. To check whether losses after selection had skewed the sample geographically, we compared the ward distribution of the analysed sample with that of the line list using a chi-square goodness-of-fit test.

### Questionnaire and measures

Trained research assistants read the questions aloud and recorded answers, since many respondents had limited schooling and the topics were personal. Interviews took place in private settings chosen so that others could not overhear. The questionnaire had sections on background characteristics; the pregnancy (whether planned, first reaction, first person told, that person’s reaction, stigma and any attempt to end the pregnancy); depressive symptoms; emotional wellbeing; and contact with support programmes.

Stigma was measured with the question “Have you been stigmatized because of your pregnancy?” (yes/no). For depressive symptoms, respondents were asked “Have you experienced any of the following since your pregnancy?” and ticked any that applied from seven items: feeling sad most days, losing interest in activities, sleeping too much or too little, poor concentration, thoughts of harming yourself, feeling hopeless about the future, and anxiety or panic attacks. An unticked item was coded as not endorsed; respondents who left the whole checklist blank were coded as missing. The items resemble those of standard screening tools, but the checklist was written for this study and has not been validated against a clinical interview. We did not use a validated instrument such as the PHQ-9, which has been validated for Kenyan adolescents [22], and our results describe self-reported symptom burden rather than the prevalence of depression. A five-item wellbeing scale (feeling supported, feeling hopeless or helpless, having someone to talk to, feeling ashamed of the pregnancy, feeling confident about the future; 1 = strongly disagree to 5 = strongly agree) was also administered. It was not used in the regression models, and its item distributions appear in Additional file 1. Respondents who had attended an advocacy or support programme on teenage pregnancy or mental health were asked who ran it, whether it included access to counselling or mental health services, and how useful they found it.

Supervisors and specialists in adolescent reproductive health reviewed the draft questionnaire for coverage, wording, suitability for minors and the handling of sensitive items on depression, stigma and abortion. It was then piloted with 21 girls (10% of the minimum sample) in Kabete Sub-County, which resembles Juja socially and demographically but lies outside the study area. A subset of pilot respondents answered the questionnaire again about two weeks later, and items that gave inconsistent or confusing answers were reworded or dropped. Internal consistency statistics were not calculated for the checklist or the wellbeing scale, because neither had been shown to measure a single underlying construct.

### Outcomes and explanatory variables

The two outcomes were stigma (yes/no) and elevated depressive symptom burden, defined before analysis as four or more of the seven checklist items (roughly the midpoint) against three or fewer. This threshold has no diagnostic standing, so we also report the unadjusted association between stigma and a lower threshold of two or more items.

Explanatory variables were chosen from the conceptual framework (Fig. 1) and not from bivariate screening. Model 1 (stigma) contained age in years at interview, school dropout (versus being in school or having completed primary or secondary school), having no income source at all (versus income from parents, a partner or own work), being single (versus married, separated or divorced; the 16 separated or divorced girls were grouped with married girls because they were too few to analyse separately), and living with parents (versus any other arrangement). Model 2 (elevated symptom burden) contained stigma, an unsupportive first reaction (anger, rejection or indifference versus support; the four girls who described a confused or worried reaction were set to missing), programme attendance, school dropout, no income source and age. Marital status was treated as a determinant of stigma and was not entered in Model 2.

### Statistical analysis

Frequencies and percentages are reported with 95% confidence intervals (CIs) from the Wilson score method [23]. Associations between categorical variables were tested with the chi-square test, applying the Yates correction to 2×2 tables and Fisher’s exact test where expected counts were small. Where a larger table had expected counts below five in more than a fifth of cells, sparse categories were merged before testing. Cramér’s V (about 0.10 small, 0.30 medium, 0.50 large) and, for 2×2 tables, unadjusted odds ratios (ORs) with 95% CIs describe the strength of association.

Both outcomes were analysed with binary logistic regression, entering all variables together. We report adjusted ORs (AORs) with 95% CIs, standard errors, Wald z values and p-values, together with the likelihood-ratio χ² for each model and McFadden’s pseudo-R². Respondents missing any model variable were excluded (complete-case analysis), leaving 199 for Model 1 and 184 for Model 2. Model 2 therefore had at most 60 outcome events for six variables, which is below the conventional ten events per variable, and its estimates should be read with this in mind [24, 25]. A planned composite measure of exposure to group counselling could not be modelled because almost all respondents fell into one category. Analyses were carried out with statistical software including IBM SPSS Statistics, version 25 [26]. Statistical significance was set at a two-sided p < .05. All results describe associations; the design does not allow causal conclusions.

### Ethical considerations

The Mount Kenya University Institutional Scientific and Ethics Review Committee approved the study (MKU/ISERC/5759; approval no. 4480). A research licence was issued by the National Commission for Science, Technology and Innovation (NACOSTI/P/26/4183417), and the County Government of Kiambu gave authorisation (KIAMBU/HRDU/PAA/2/2026). Girls aged 18 years and above gave written informed consent. For girls under 18, a parent or guardian gave written consent and the girl gave written assent. Participants were told they could withdraw at any time without any effect on the services they received. Data were handled under Kenya’s Data Protection Act, 2019 [27]: names were not entered in the analysis file, each respondent had a code, and the list linking codes to names was kept apart and was available only to the principal investigator. Before fieldwork began, we agreed a referral arrangement with local health facilities. Any girl who reported active thoughts of suicide, or who appeared acutely distressed, was linked with her agreement to a named facility contact for psychosocial support on the same day.

## Results

### Characteristics of respondents

Figure 2 shows the flow of participants. The line list of 466 girls was the sampling frame, and 227 girls (48.7%) contributed a valid questionnaire to the analysis. The remaining 239 girls on the list were not part of the analysed sample. They include girls who were never selected, because the design required only part of the frame (minimum 211), and selected girls who declined, were not interviewed because of acute distress, or whose questionnaires were incomplete or invalid at data cleaning. These groups were not counted separately, so we cannot report a response rate. The analysed sample nevertheless mirrored the frame closely: each ward contributed between 48.5% and 50.0% of its listed girls, and the ward distribution of the sample did not differ from that of the line list (χ²(4) = 0.02, p > .99). Item-level missing data reduced the number available for each analysis: 226 girls completed the symptom checklist, 221 had both checklist and stigma data, and 199 and 184 had complete data for Models 1 and 2.

**Fig. 2.**
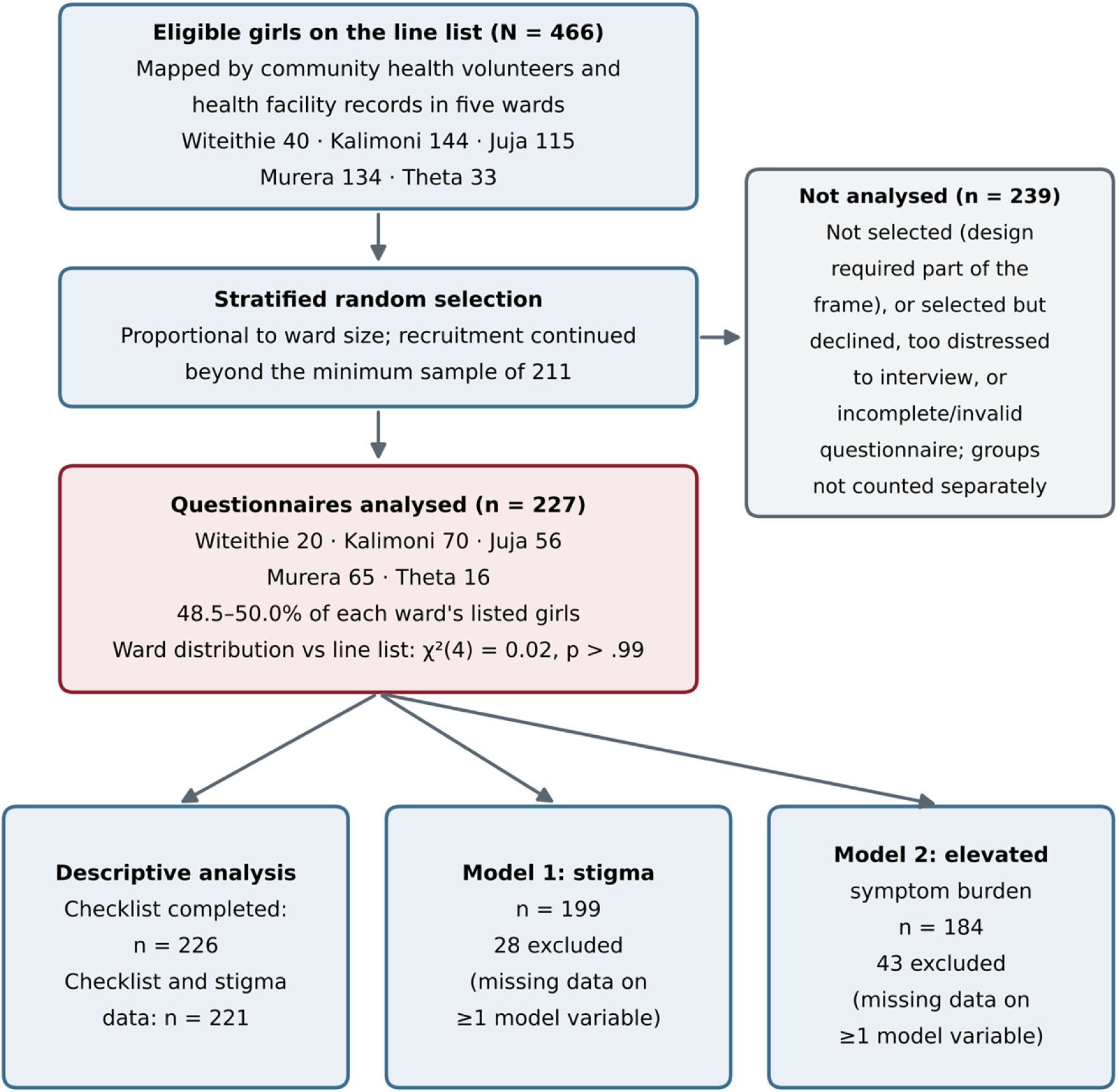
Flow of participants through the study, from the community line list to the analytic samples for the descriptive analysis and the two regression models.

Most respondents were 17–19 years old (86.8%), and 19-year-olds alone made up 38.3% of the sample. Parity was recorded for 197 girls, and 194 of them (98.5%) had already given birth, nearly all to one child; three were pregnant for the first time. The sample is therefore mainly one of young mothers. Almost half (47.1%) had left school before completing it, and one in ten was still enrolled. Seven in ten were single, about two-thirds lived with their parents or a guardian, and only 13 (5.7%) had their own job or other income; the rest had no income (38.8%) or relied on parents (36.6%) or a partner (17.2%). None of the 224 girls who answered the question reported a disability or chronic illness (Table 1).

### Pregnancy, disclosure, stigma and programme contact

Nine in ten pregnancies (91.2%) had not been planned. On learning they were pregnant, 40.1% of girls felt confused, 26.4% afraid and 21.6% depressed; 9.3% felt happy. The first person told was most often the girl’s mother (32.6%), then her partner (22.5%) or a friend (21.1%), and 17.2% initially told no one. Half of first disclosures (50.2%) met with support. Anger (20.7%), rejection (13.7%) and indifference (7.0%) together accounted for 41.4%. Stigma was reported by 43.2% of girls, and 13.2% had tried to end the pregnancy, mostly at home (10.6%) rather than at a facility (2.6%) (Table 2).

**Table 2.**
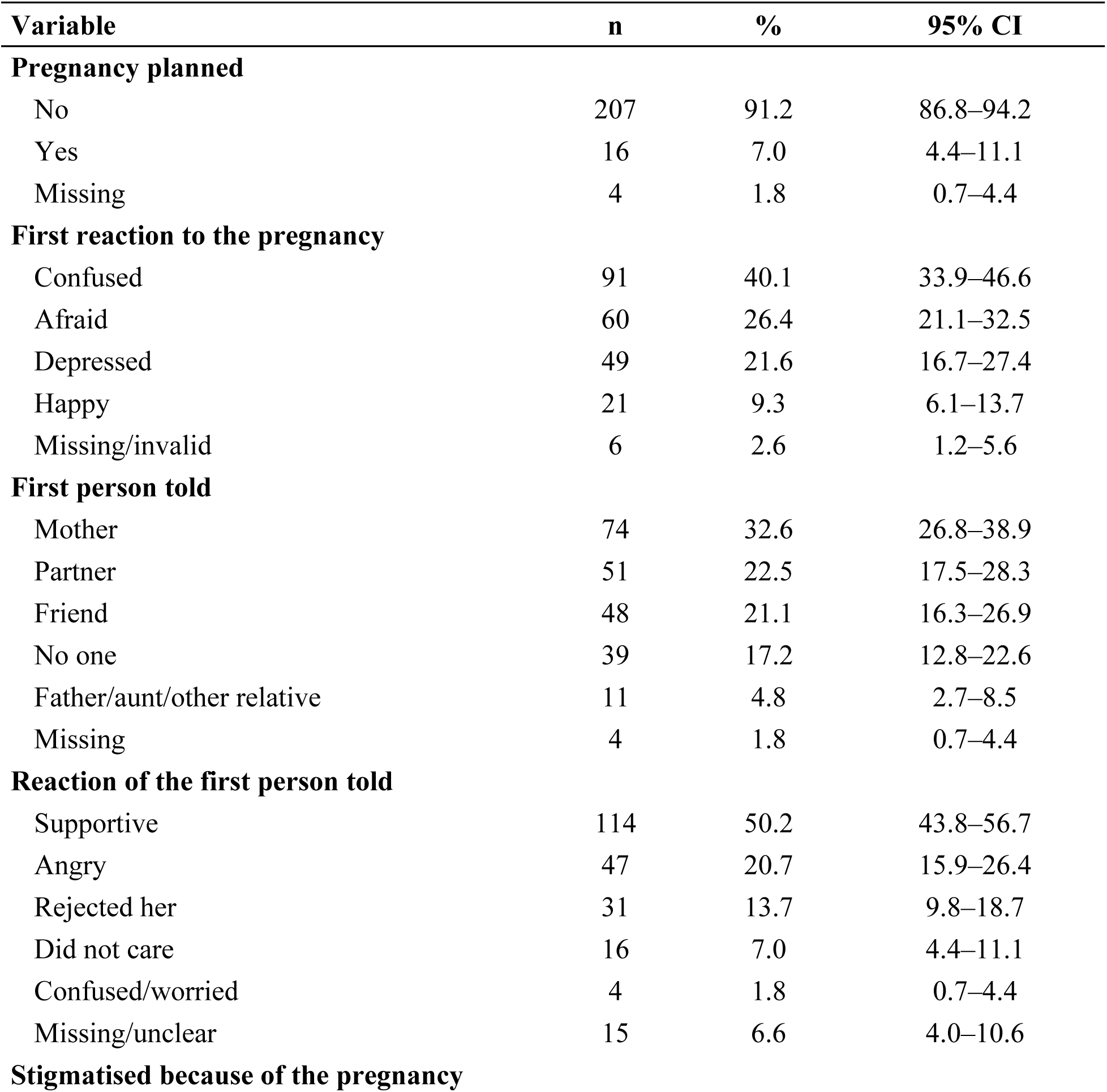
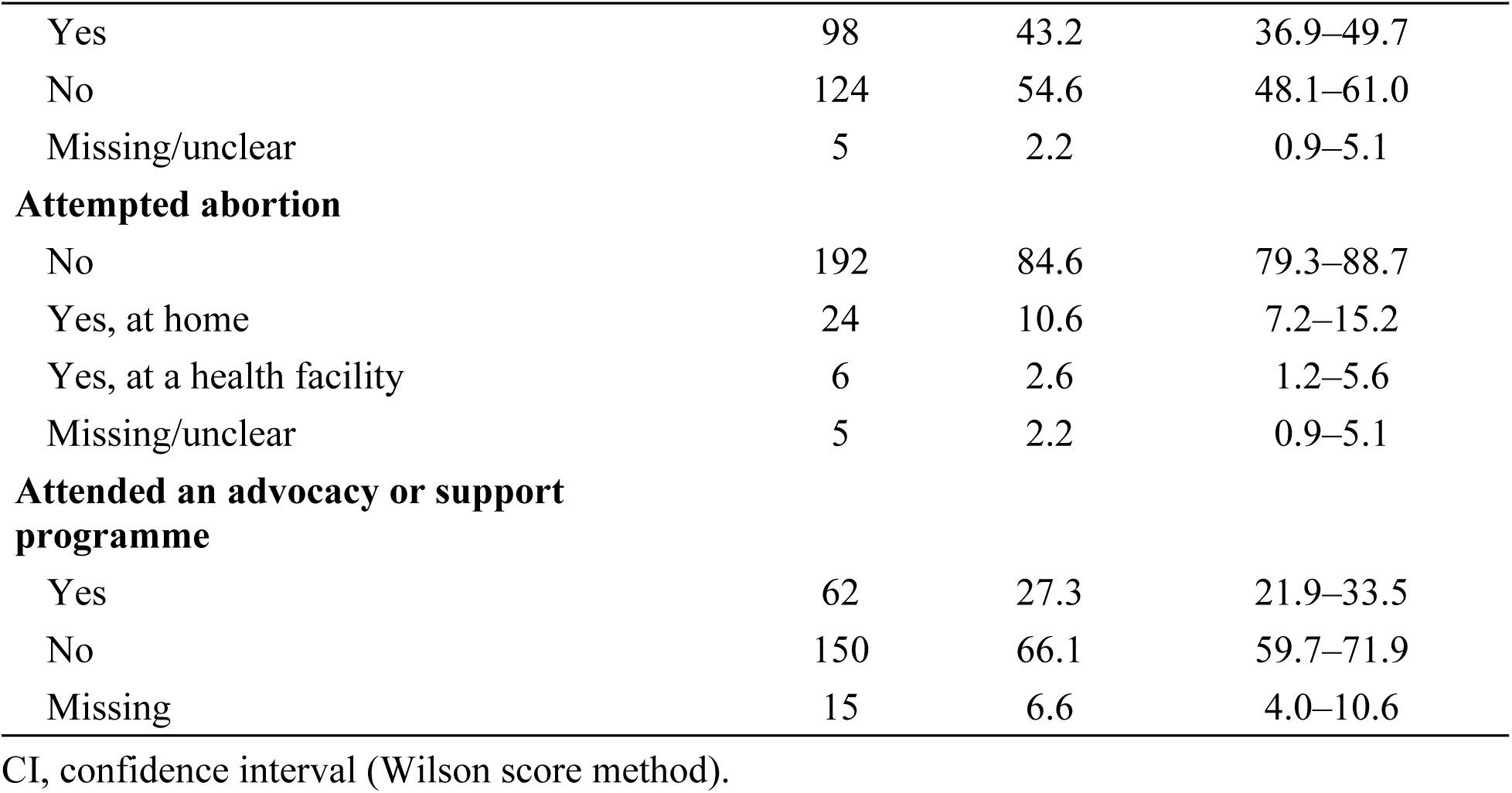
Pregnancy, disclosure, stigma and programme contact (N = 227)

| **Variable** | **n** | **%** | **95% CI** |
|---|---|---|---|
| **Pregnancy planned** |  |  |  |
| No | 207 | 91.2 | 86.8–94.2 |
| Yes | 16 | 7.0 | 4.4–11.1 |
| Missing | 4 | 1.8 | 0.7–4.4 |
| **First reaction to the pregnancy** |  |  |  |
| Confused | 91 | 40.1 | 33.9–46.6 |
| Afraid | 60 | 26.4 | 21.1–32.5 |
| Depressed | 49 | 21.6 | 16.7–27.4 |
| Happy | 21 | 9.3 | 6.1–13.7 |
| Missing/invalid | 6 | 2.6 | 1.2–5.6 |
| **First person told** |  |  |  |
| Mother | 74 | 32.6 | 26.8–38.9 |
| Partner | 51 | 22.5 | 17.5–28.3 |
| Friend | 48 | 21.1 | 16.3–26.9 |
| No one | 39 | 17.2 | 12.8–22.6 |
| Father/aunt/other relative | 11 | 4.8 | 2.7–8.5 |
| Missing | 4 | 1.8 | 0.7–4.4 |
| **Reaction of the first person told** |  |  |  |
| Supportive | 114 | 50.2 | 43.8–56.7 |
| Angry | 47 | 20.7 | 15.9–26.4 |
| Rejected her | 31 | 13.7 | 9.8–18.7 |
| Did not care | 16 | 7.0 | 4.4–11.1 |
| Confused/worried | 4 | 1.8 | 0.7–4.4 |
| Missing/unclear | 15 | 6.6 | 4.0–10.6 |
| **Stigmatised because of the pregnancy** |  |  |  |
| Yes | 98 | 43.2 | 36.9–49.7 |
| No | 124 | 54.6 | 48.1–61.0 |
| Missing/unclear | 5 | 2.2 | 0.9–5.1 |
| **Attempted abortion** |  |  |  |
| No | 192 | 84.6 | 79.3–88.7 |
| Yes, at home | 24 | 10.6 | 7.2–15.2 |
| Yes, at a health facility | 6 | 2.6 | 1.2–5.6 |
| Missing/unclear | 5 | 2.2 | 0.9–5.1 |
| **Attended an advocacy or support programme** |  |  |  |
| Yes | 62 | 27.3 | 21.9–33.5 |
| No | 150 | 66.1 | 59.7–71.9 |
| Missing | 15 | 6.6 | 4.0–10.6 |

CI, confidence interval (Wilson score method).

Only 62 girls (27.3% of the sample; 29.2% of the 212 who answered) had attended any advocacy or support programme. Of the 59 attendees who answered the follow-up question, 41 (69.5%) said the programme included access to counselling or mental health services, so about 18% of the whole sample had contact with a programme offering counselling. Of 60 attendees who rated the programme, 42 (70.0%) found it very useful and 12 (20.0%) somewhat useful.

How the first confidant reacted depended on who that person was (χ²(6, n = 197) = 36.80, p < .001; Cramér’s V = 0.31; rejection and indifference were merged for this test so that every expected count was at least five). Partners responded with support in 37 of 50 cases (74.0%) and mothers in 42 of 70 (60.0%), while 22 of the 70 mothers (31.4%) reacted with anger. The 30 girls who told no one, and whose pregnancies others presumably discovered, fared worst. Six of them (20.0%) eventually received support, twelve (40.0%) faced anger and seven (23.3%) faced indifference (Table 3). Girls who confided first in a partner were probably more often married or cohabiting, so the higher rate of partner support may partly reflect marital status.

**Table 3.**
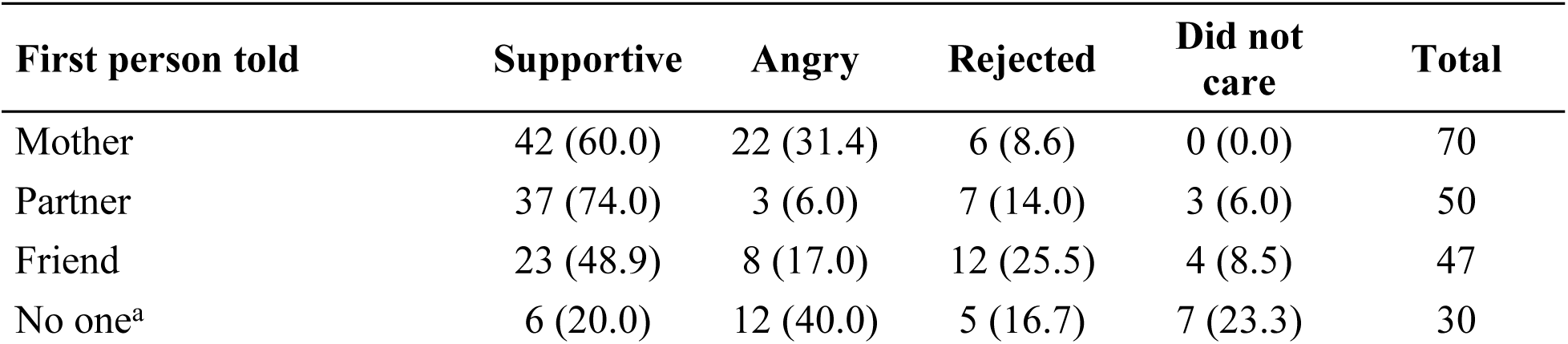
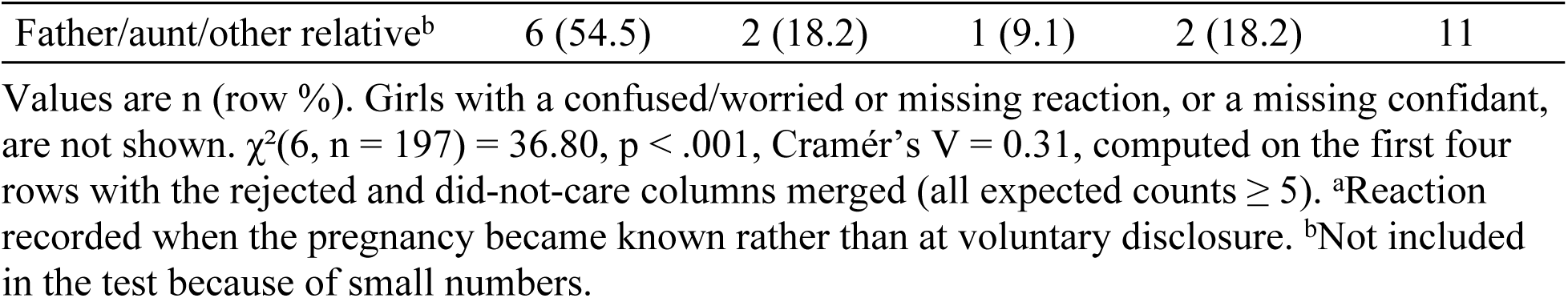
Reaction received, by first person told.

| **First person told** | **Supportive** | **Angry** | **Rejected** | **Did not care** | **Total** |
|---|---|---|---|---|---|
| Mother | 42 (60.0) | 22 (31.4) | 6 (8.6) | 0 (0.0) | 70 |
| Partner | 37 (74.0) | 3 (6.0) | 7 (14.0) | 3 (6.0) | 50 |
| Friend | 23 (48.9) | 8 (17.0) | 12 (25.5) | 4 (8.5) | 47 |
| No one<sup>a</sup> | 6 (20.0) | 12 (40.0) | 5 (16.7) | 7 (23.3) | 30 |
| Father/aunt/other relative<sup>b</sup> | 6 (54.5) | 2 (18.2) | 1 (9.1) | 2 (18.2) | 11 |

Values are n (row %). Girls with a confused/worried or missing reaction, or a missing confidant, are not shown. χ<sup>2</sup>(6, n = 197) = 36.80, p < .001, Cramér’s V = 0.31, computed on the first four rows with the rejected and did-not-care columns merged (all expected counts ≥ 5). <sup>a</sup>Reaction recorded when the pregnancy became known rather than at voluntary disclosure. <sup>b</sup>Not included in the test because of small numbers.

### Depressive symptoms

Of the 226 girls who completed the checklist, 62.8% reported disturbed sleep, 50.4% loss of interest and 50.0% feeling sad most days. Hopelessness about the future (35.8%), poor concentration (32.3%) and anxiety or panic attacks (24.8%) were less common. Thirty-seven girls (16.4%) reported thoughts of harming themselves (Table 4, Fig. 3). Among the 221 girls with both checklist and stigma data, 75 (33.9%) endorsed none or one item, 86 (38.9%) two or three items, and 60 (27.1%; 95% CI 21.7–33.4) four or more items, meeting our definition of elevated symptom burden. Answers on the wellbeing scale clustered at the two ends of the range (Additional file 1).

**Fig. 3.**
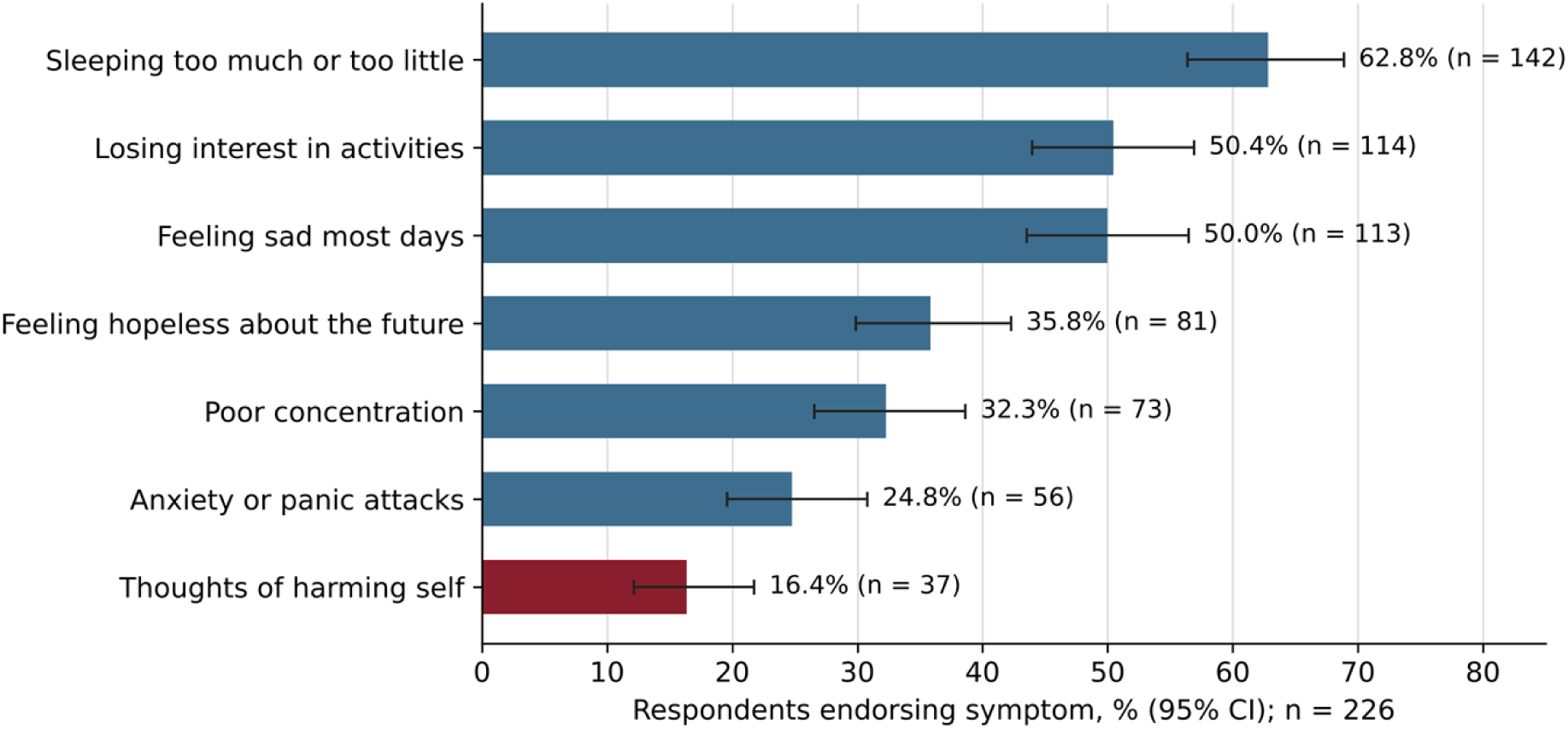
Proportion of respondents endorsing each checklist item, with 95% Wilson confidence intervals (n = 226). The self-harm item is shown in red.

**Table 4.**
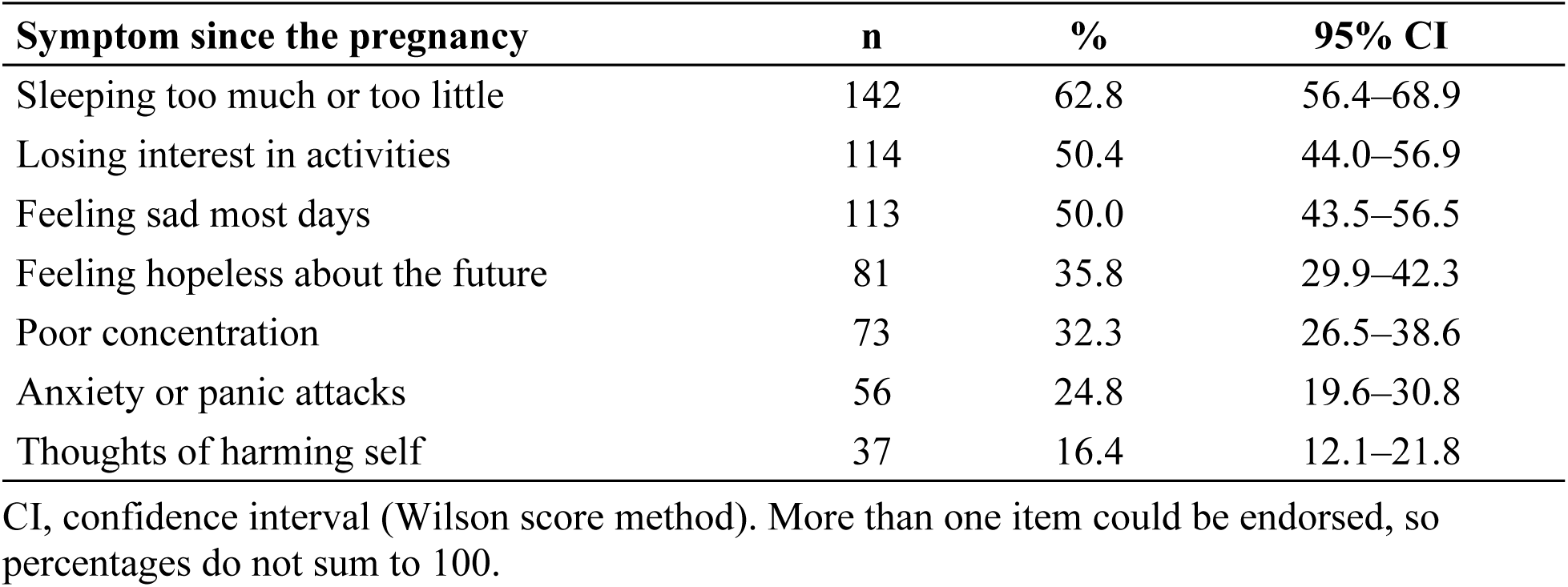
Endorsement of depressive symptom checklist items (n = 226)

### Unadjusted associations with stigma

Table 5 compares girls who did and did not report stigma. Stigma did not differ by schooling (χ²(3) = 3.36, p = .339), although it was somewhat more common among girls who had dropped out (50.5%) than among those who had completed secondary school (35.7%; unadjusted OR for dropout versus all others 1.56, 95% CI 0.91–2.68). Stigma varied with the first reaction to the pregnancy (χ²(3) = 20.12, p < .001; V = 0.30). It was reported by 59.2% of girls whose first reaction was feeling depressed, but by only one of the 21 girls (4.8%) who had felt happy. Girls who had attempted an abortion were more often stigmatised than those who had not (65.5% vs 41.4%; OR 2.69, 95% CI 1.19–6.10; p = .025). Symptom burden rose with stigma: 68.3% of girls with four or more symptoms reported stigma, against 30.7% of those with none or one (χ²(2) = 20.95, p < .001; V = 0.31). Put the other way round, 42.3% of stigmatised girls and 15.3% of the others had elevated symptom burden (OR 4.05, 95% CI 2.15–7.62). The association held at a lower threshold of two or more symptoms (76.3% vs 58.1%; OR 2.32, 95% CI 1.29–4.19). Programme attenders reported stigma more often than non-attenders (57.4% vs 38.0%; OR 2.20, 95% CI 1.20–4.02; p = .016). Programmes may have drawn in, or been referred, girls in greatest difficulty, or attendance may have made girls more visible to their neighbours; these data cannot tell the two apart.

**Table 5.** Stigma by selected characteristics.

| **Characteristic** | **n** | **Stigmatised, n (%)** | **χ² (df)** | **p** | **Cramér’s V** |
|---|---|---|---|---|---|
| **Schooling status** |  |  |  |  |  |
| Dropped out | 107 | 54 (50.5) | 3.36 (3) | .339 | 0.12 |
| Completed secondary | 56 | 20 (35.7) |  |  |  |
| Completed primary | 31 | 13 (41.9) |  |  |  |
| In school | 22 | 10 (45.5) |  |  |  |
| **First reaction to the pregnancy** |  |  |  |  |  |
| Confused | 90 | 35 (38.9) | 20.12 (3) | <.001 | 0.30 |
| Afraid | 60 | 31 (51.7) |  |  |  |
| Depressed | 49 | 29 (59.2) |  |  |  |
| Happy | 21 | 1 (4.8) |  |  |  |
| **Attempted abortion**<sup>a</sup> |  |  |  |  |  |
| Yes (home or facility) | 29 | 19 (65.5) | 5.01 (1) | .025 | 0.15 |
| No | 191 | 79 (41.4) |  |  |  |
| **Attended a support programme**<sup>a</sup> |  |  |  |  |  |
| Yes | 61 | 35 (57.4) | 5.86 (1) | .016 | 0.17 |
| No | 150 | 57 (38.0) |  |  |  |
| **Depressive symptoms endorsed** |  |  |  |  |  |
| 0–1 | 75 | 23 (30.7) | 20.95 (2) | <.001 | 0.31 |
| 2–3 | 86 | 33 (38.4) |  |  |  |
| 4–7 | 60 | 41 (68.3) |  |  |  |

Row percentages. Girls missing either variable are excluded, so totals differ between sections; for schooling, the six girls who had never attended school or whose status was unclear are excluded. <sup>a</sup>Yates continuity correction applied.

### Factors associated with stigma

Model 1 (n = 199) was statistically significant (likelihood-ratio χ²(5) = 20.96, p = .001), but its pseudo-R² of 0.076 shows that the five variables explained only a small part of the variation in stigma. Being single was the only significant factor. Single girls had 3.50 times the odds of reporting stigma of girls who were married, separated or divorced, after adjustment for age, schooling, income and living arrangement (95% CI 1.48–8.31; p = .004). Dropout, lack of income, living with parents and age were not associated with stigma in the adjusted model (Table 6, Fig. 4).

**Fig. 4.**
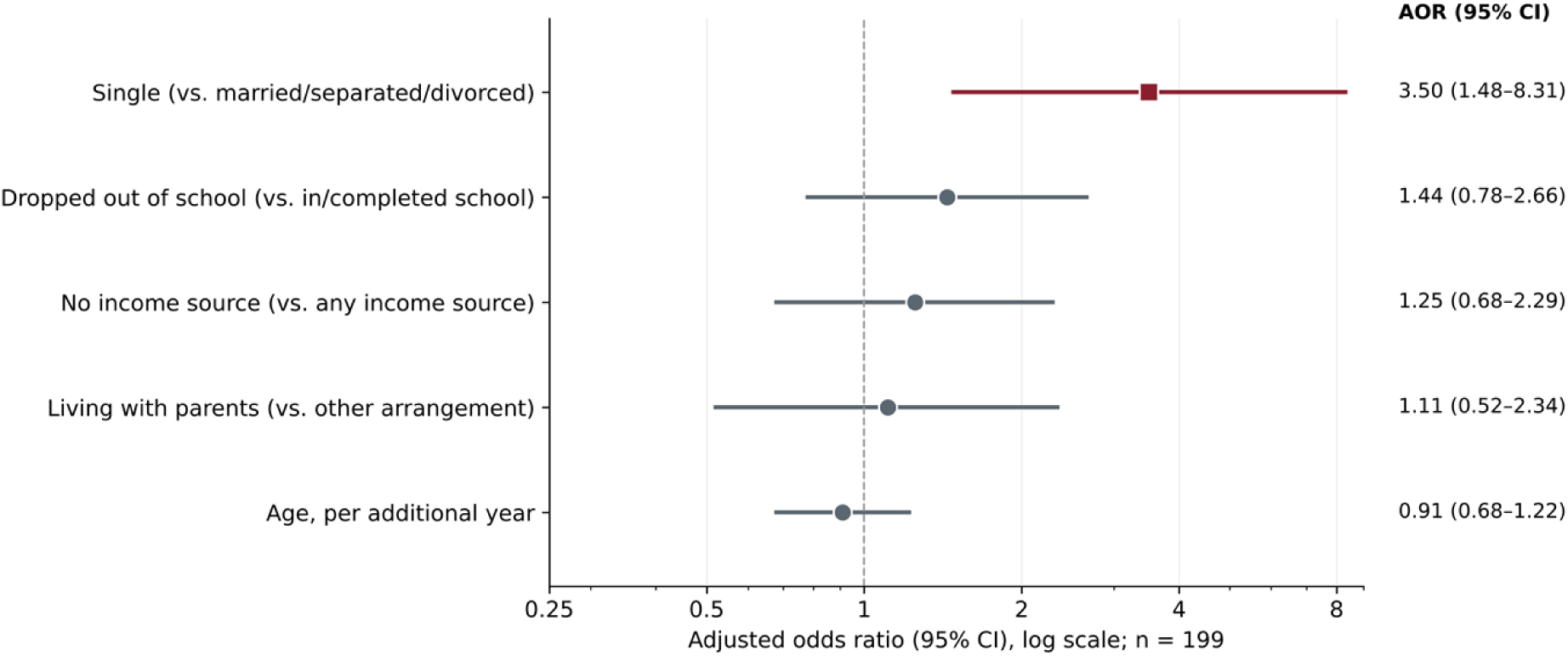
Adjusted odds ratios (95% CI) for factors associated with stigma (Model 1, n = 199). Red squares, p < .05; grey circles, p ≥ .05; dashed line, AOR = 1.

**Table 6.** Factors associated with stigma: logistic regression (Model 1, n = 199)

| Variable | AOR | 95% CI | SE | z | p |
|---|---|---|---|---|---|
| Single (vs. married/separated/divorced) | 3.50 | 1.48–8.31 | 0.44 | 2.84 | .004 |
| Dropped out of school (vs. in/completed school) | 1.44 | 0.78–2.66 | 0.31 | 1.17 | .243 |
| No income source (vs. any income) | 1.25 | 0.68–2.29 | 0.31 | 0.71 | .477 |
| Living with parents (vs. other arrangement) | 1.11 | 0.52–2.34 | 0.38 | 0.26 | .792 |
| Age, per additional year | 0.91 | 0.68–1.22 | 0.15 | −0.63 | .528 |

AOR, adjusted odds ratio; CI, confidence interval; SE, standard error of the log odds; z, Wald statistic. Likelihood-ratio χ²(5) = 20.96, p = .001; McFadden’s pseudo-R² = 0.076.

### Factors associated with elevated depressive symptom burden

Model 2 (n = 184) explained more of the variation in its outcome than Model 1 (likelihood-ratio χ²(6) = 35.82, p < .001; McFadden’s pseudo-R² = 0.162). With the other variables held constant, the odds of elevated symptom burden were 3.21 times higher among girls who reported stigma (95% CI 1.54–6.69; p = .002), and girls whose first confidant reacted with anger, rejection or indifference had 2.54 times the odds (95% CI 1.19–5.41; p = .016). The odds fell by 31% with each additional year of age (AOR 0.69, 95% CI 0.49–0.96; p = .030). Having no income source was associated with higher odds, but the estimate was imprecise and not statistically significant (AOR 1.90, 95% CI 0.92–3.94; p = .084). School dropout (AOR 0.99; p = .987) and programme attendance (AOR 0.89; p = .771) showed no association (Table 7, Fig. 5).

**Fig. 5.**
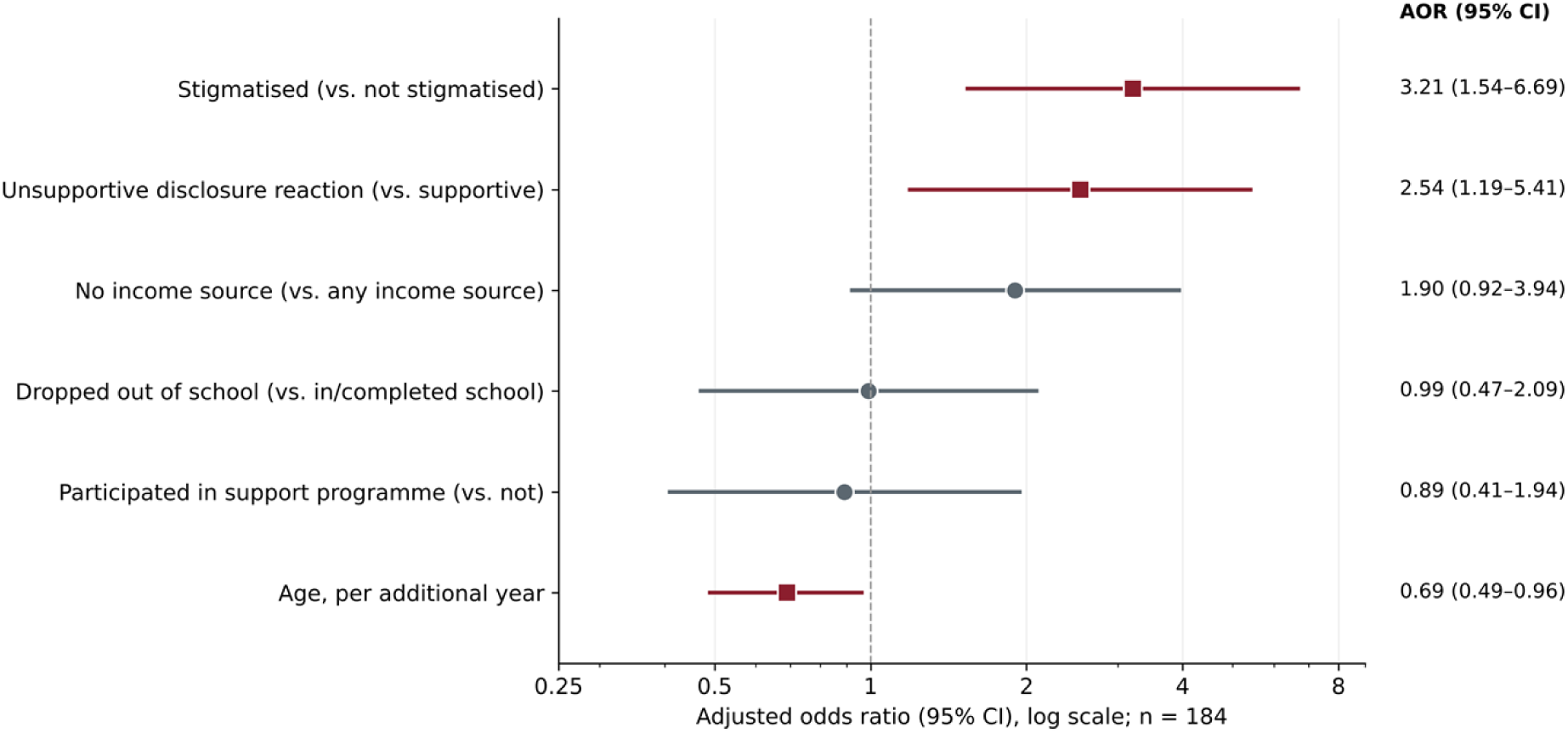
Adjusted odds ratios (95% CI) for factors associated with elevated depressive symptom burden (Model 2, n = 184). Red squares, p < .05; grey circles, p ≥ .05; dashed line, AOR = 1.

**Table 7.** Factors associated with elevated depressive symptom burden: logistic regression (Model 2, n = 184)

| **Variable** | **AOR** | **95% CI** | **SE** | **z** | **p** |
|---|---|---|---|---|---|
| Stigmatised (vs. not) | 3.21 | 1.54–6.69 | 0.37 | 3.11 | .002 |
| Unsupportive first reaction (vs. supportive) | 2.54 | 1.19–5.41 | 0.39 | 2.42 | .016 |
| No income source (vs. any income) | 1.90 | 0.92–3.94 | 0.37 | 1.73 | .084 |
| Dropped out of school (vs. in/completed school) | 0.99 | 0.47–2.09 | 0.38 | −0.02 | .987 |
| Attended a support programme (vs. not) | 0.89 | 0.41–1.94 | 0.40 | −0.29 | .771 |
| Age, per additional year | 0.69 | 0.49–0.96 | 0.17 | −2.17 | .030 |

Outcome: four or more of seven checklist items vs. three or fewer. AOR, adjusted odds ratio; CI, confidence interval; SE, standard error of the log odds; z, Wald statistic. Likelihood-ratio χ<sup>2</sup>(6) = 35.82, p < .001; McFadden’s pseudo-R<sup>2</sup> = 0.162.

## Discussion

In this community sample of mostly young mothers, about a quarter had elevated depressive symptom burden and one in six reported thoughts of self-harm. Symptoms were concentrated among girls who had been stigmatised and among those whose first confidant reacted badly, and each of these associations remained after the other was taken into account, along with income, schooling, programme attendance and age. Stigma itself was linked to being unmarried, not to schooling, income or living arrangement. These results fit with earlier work on stigma and depressive symptoms in pregnant adolescents in Nairobi and Mwanza [8–10], and extend it to a community sample of girls who have already given birth.

The adjusted OR for stigma (3.21) is larger than the figure of about 1.8 reported for pregnancy-related stigma among pregnant adolescents attending antenatal clinics in Mwanza [10]. Measurement is one likely reason: we used a single global stigma question and an unvalidated checklist, whereas the Mwanza team used the Edinburgh Postnatal Depression Scale. The populations also differ. Our respondents were mostly past the birth, many had left school, and they were found through community lists rather than at a clinic. Stigma may be more visible, and less softened by contact with services, in that situation. The link between younger age and more symptoms matches the Kangemi finding that the youngest pregnant adolescents were most at risk [9], and the observation from Cape Town that girls under 17 expressed more fear and shame [16]. Our age variable was current age, and we could not separate it from time since birth because almost no respondent answered the question on age at pregnancy, so this result needs cautious reading.

The association of stigma with marital status rather than with poverty or schooling is what Link and Phelan’s account would predict [13]: the label attaches to childbearing outside marriage, not to being poor. The same pattern has been described among unmarried adolescent mothers in Nigeria [15], in the moral framing of adolescent pregnancy in Cape Town [16], and in Kenyan nurses’ explanations of how they treat pregnant teenagers [17]. If the pattern holds more widely, programmes that offer only economic help, or that call in general terms for acceptance, will leave the normative basis of stigma untouched. The low pseudo-R² of Model 1, however, shows that most of the variation in stigma came from things we did not measure.

An unsupportive first reaction was associated with more symptoms independently of stigma, which is in line with the main-effect model of social support [14]. We did not test whether a supportive reaction weakens the link between stigma and symptoms; that question needs an interaction term and a larger sample. Partners were the most consistently supportive first confidants. Many of these girls were probably married or living with the partner, so the finding is likely confounded by marital status, and it is not evidence that telling a partner is always safe. Among adolescent mothers in urban South Africa, partner violence during pregnancy was associated with a higher risk of postpartum depressive symptoms (adjusted risk ratio 1.62) [28], and we did not ask about partner violence.

The 16.4% of girls who reported thoughts of harming themselves is the most urgent result. Data on self-harm and suicidal behaviour among pregnant and parenting adolescents in sub-Saharan Africa are scarce, and researchers have called for direct measurement [3, 29]. Our item asked about thoughts of self-harm and is not the same as a measure of suicidal ideation. Even so, the figure, set beside the finding that fewer than three in ten girls had attended any programme and fewer than one in five had attended one offering counselling, supports routine screening wherever these girls already come into contact with services.

Programme attendance was not associated with fewer symptoms, and stigmatised girls were more likely to have attended. A yes/no attendance question says nothing about what a programme contained or how long it lasted, and with cross-sectional data the direction of the link is unknown. Attenders mostly rated their programme as useful, so the problem may be one of reach and content rather than of how programmes are received. A systematic review found little evidence that psychosocial interventions reduce depression or anxiety in pregnant and parenting adolescents, and none of the included trials took place in a low- or middle-income country [30]. Group interpersonal psychotherapy delivered by trained lay workers reduced depression in Uganda [31] and is being adapted for pregnant and parenting adolescents in Kenya [32, 33]; it is one candidate for addressing the factors identified here.

### Strengths and limitations

We sampled from community lists, which allowed us to reach girls who had left school or were not attending antenatal care, groups that Kenyan clinic studies miss. Stigma, disclosure experience and economic factors were analysed together in models specified from theory, and we report effect sizes and confidence intervals throughout.

The study also has important limitations. Because the design is cross-sectional, we cannot say whether stigma and poor disclosure reactions came before depressive symptoms; girls with more symptoms may also be more likely to perceive or report stigma, or to remember a reaction as hostile. The checklist has not been validated, the four-item threshold has no clinical basis, and the results cannot be compared directly with PHQ-9 or EPDS figures. Its tick-all-that-apply format means that an item left blank was counted as absent, which could understate symptoms. Sleep disturbance, the most common item, may partly reflect caring for an infant. Stigma was measured with one question that does not distinguish experienced, anticipated and internalised stigma or say where it came from. Time since birth was not recorded, and the age-at-pregnancy question was almost never answered, so the reaction to disclosure may have been recalled after a long and variable interval. Parental consent was required for girls under 18, and highly distressed girls were not interviewed; both may have excluded those with the most hostile families or the most severe symptoms. The community health volunteer lists may have missed girls unknown to local services. Selected girls who declined, were too distressed to interview or returned incomplete questionnaires were not counted separately from girls who were never selected, so no response rate can be given. The close match between the ward distribution of the sample and the line list argues against large losses in any one area, but losses related to individual characteristics, such as family hostility or severe distress, cannot be ruled out. Complete-case analysis reduced the sample to 199 and 184, and Model 2 had fewer than ten events per variable. Marital status was not included in Model 2, so it may confound the association between stigma and symptoms. We did not measure partner violence, sexual coercion, HIV status or previous mental health problems, which is consistent with the modest explanatory power of both models. No girl reported a disability, which more likely reflects the wording of the question or reluctance to answer than a true absence. Finally, the findings come from one peri-urban sub-county and may not apply where social norms differ.

### Implications

Brief screening with a validated tool such as the PHQ-9 [22], with direct questions on self-harm and suicidal thoughts and a clear route to care, could be built into postnatal, child-health and immunisation visits, which reach adolescent mothers after birth, and into antenatal care, which most Kenyan women attend (two in three make four or more visits) [5]. Families and partners could be helped to respond well when a girl discloses a pregnancy. Stigma-reduction work could address attitudes to unmarried motherhood directly. Each of these needs evaluation. Future studies should use validated depression measures and multidimensional stigma scales, record time since birth, ask about partner violence and follow girls over time so that the order of events can be established.

## Conclusions

Elevated depressive symptom burden was common among adolescent mothers in Juja Sub-County and was concentrated among girls who reported stigma or who met anger, rejection or indifference when they first disclosed their pregnancy. Stigma was linked to being unmarried rather than to economic hardship, which suggests that economic support or general awareness campaigns alone will not be enough. Mental health screening with validated tools at postnatal, child-health and antenatal visits, support for families and partners at the point of disclosure, and community work on attitudes to unmarried motherhood should be tested as priorities in this and similar Kenyan settings.

## Data Availability

The dataset analysed in this study is available from the corresponding authors on reasonable request, subject to the ethical and data-protection conditions under which it was collected.

## Abbreviations

AOR: adjusted odds ratio
CI: confidence interval
EPDS: Edinburgh Postnatal Depression Scale
NACOSTI: National Commission for Science, Technology and Innovation
OR: odds ratio
PHQ-9: Patient Health Questionnaire-9
SE: standard error
STROBE: Strengthening the Reporting of Observational Studies in Epidemiology.

## Declarations

### Ethics approval and consent to participate

The study was approved by the Mount Kenya University Institutional Scientific and Ethics Review Committee (MKU/ISERC/5759; approval no. 4480), licensed by the National Commission for Science, Technology and Innovation (NACOSTI/P/26/4183417) and authorised by the County Government of Kiambu (KIAMBU/HRDU/PAA/2/2026). Participants aged 18 years and above gave written informed consent. For participants under 18 years, a parent or guardian gave written consent and the participant gave written assent. The study was conducted in accordance with the Declaration of Helsinki.

### Consent for publication

Not applicable.

### Authors’ contributions

TWK designed the study, led data collection and analysis, and wrote the first draft with guidance from GM. JK and DM supervised the study. JK, DM, JO and GM contributed to the design and analysis and revised the manuscript critically. All authors read and approved the final manuscript.

### Clinical trial number

Not applicable.

## Acknowledgements

We thank the young women who took part, the community health volunteers who helped with mapping and recruitment, and the research assistants who conducted the interviews.

## Use of artificial intelligence tools

The authors used Grammarly (Grammarly Inc.) to refine language, grammar and readability. All content was written, reviewed and approved by the authors, who take full responsibility for this article. No AI tool is listed as an author.

## Supporting information

**S1 Checklist. STROBE checklist for cross-sectional studies.** Location in the manuscript of each item of the STROBE statement.

**S1 Table. Responses to the emotional wellbeing scale.** Distribution of responses to the five wellbeing items, with Wilson 95% confidence intervals.

